# Acute Kidney Injury, the Present on Admission indicator (POA) and sex disparities: Observational study of inpatient real world data in a Swiss tertiary health care system

**DOI:** 10.1101/2023.01.16.23284622

**Authors:** Karen Triep, Sarah Musy, Michael Simon, Olga Endrich

**Affiliations:** Medical Directorate, Inselspital, University Hospital of Bern, Berne, Switzerland; Institute of Nursing Science, Department Public Health, Faculty of Medicine, University of Basel, Basel, Switzerland; Center for Laboratory Medicine (ZLM)/University Institute of Clinical Chemistry, Inselspital, University Hospital, University of Bern, Berne, Switzerland

## Abstract

**Background:** Regarding kidney disease, sex differences in epidemiology and clinical relevance have been reported. Related to absolute and relative changes of baseline creatinine, different criteria for staging may induce under-or over-diagnosis related to sex. At the largest Swiss provider of inpatient acute health care, a clinic decision support algorithm ensures exact staging of kidney disease (2012 KDIGO Clinical Practice Guideline). Coding of the indicator “Present On Admission” was introduced at this institution in 2018 to flag post-admission conditions.

**Objective:** We hypothesized sex differences in health care associated acute kidney injury. Defined indicators and the distribution of stages in acute kidney injury were analysed using the POA flag. Sex differences were reported.

**Methods:** Retrospective observational study. Routinely collected health data, Insel Group, Berne, Switzerland, 2019 and 2020 (121’757 cases) on the patient history and intensive care treatment duration, comorbidity levels, coded diagnoses, age and sex. Software and statistic: program R, version 4.1.1, standard deviation; median, interquartile range; prop.test; standardized mean difference.

**Results:** The reporting of post-admission diagnoses was associated with more interhospital transfers, intensive care stays, scores of severity and treatment intensity, mechanical ventilation, age, number of diagnoses, complexity level of the related cases and mortality. A weaker association could be observed for the female population. However, mortality was higher (stage III acute kidney injury 41.6%).

**Conclusion:** Using the POA-flag the results reflect the clinical situation of complications and comorbidities evolving unexpectedly. As our results show sex differences, i.e. a lower morbidity of female patients for each stage, but a higher mortality, a deeper evaluation of the implied sex differences in staging of kidney disease should follow.

The general results confirm the necessity of a diagnosis-onset reporting in health statistic.

## Introduction

In many studies, it is assumed that data obtained from research involving male participants could be extrapolated to females (1–5). However, the epidemiology and outcome of many diagnoses are related to biological sex (6). Sex has shown to influence medical diagnoses (7,8). Not only the biological but also other different determinants from social and economic to individual behaviours of patients and providers possibly contribute to the reported differences (9). The sex bias which is still present in the majority of research disciplines prefers male subjects despite research policies, e.g. the European research programme Horizon 2020 (7)(10).

Kidney disease (KD) is a particularly relevant population with sex differences. There are more women with chronic kidney disease (CKD), yet only 40% of patients receiving kidney replacement therapy are female (11–14). Disparities in access to care are described for Intensive Care Unit (ICU) admission, less aggressive medical care and later initiation of renal replacement therapy (15–17). Even in high income countries, the access to kidney transplantation seems to be limited for women and they receive less and donate more organs (11,16). The lacking treatment corresponds to outcomes of CKD as well as of acute kidney injury (AKI) by increasing the acuity, incidence and severity (6,13,14,16,18). Relevant underrepresentation of female participants could be made transparent by a recent study focussing on HIV/AIDS, CKD, and cardiovascular diseases among others (10). The underrepresentation of female sex applies not only for study populations involving disease diagnosis but also several scoring systems, like severity and nursing scores for intensive care treatment (ICU; TISS-28, SAPS, NEMS) (19–21).

Biological sex is increasingly recognized as modulator of the pathophysiology, course and progression of disease and management of CKD. This also has been examined in hospital acquired AKI (HA-AKI) (13,22–24). The introduction of the KDIGO guidelines (2012 KDIGO Clinical Practice Guideline for AKI and the Clinical Practice Guideline for the Evaluation and Management of CKD (25,26)) to assess AKI and CKD raised questions concerning sex differences due to definition criteria (23,24,27). Studies with unadjusted risk stratification found that indipendently from diagnosis criteria used males were significantly more likely to develop HA-AKI than females (27). Whatever the concluding discussion will convey in the future there is no doubt that for both sex the prevalences are high for both AKI and CKD, with AKI ranging from 10.7% to 31.3% (12,28,29), that sex differences in epidemiology and in the transition from AKI to CKD can be observed (22,30) and that HA-AKI is associated with increased mortality and resource consumption (31–35). Moreover, for both sex AKI, CKD, and acute-on-chronic KD are very relevant as a comorbidity, intercurrent disease, or complication (28,36,37).

Indexing clinical information to sex and diagnosis-timing, i.e. using a present on admission flag for hospital admission is important in order to monitor complications, intercurrent diseases and disease progression in CKD and AKI. The use of routinely collected health data of inpatient care for understanding patterns of different outcomes has been limited in Switzerland by the fact that pre-existing and post-admission conditions have been indistinguishable. In Switzerland, encoded data of inpatient stays are submitted to the Federal Office of Statistics (BFS) for publication of epidemiological and economic health care statistics (38). Furthermore, they are used for pricing and reimbursement according to Swiss Diagnosis Related Groups (Swiss DRG) (39). The Swiss national medical statistics does not provide information on the timing of the diagnosis. The use of a “Present on Admission Indicator” (POA) allows to differentiate diagnoses which arise during from those arising before an inpatient stay and therefore provide an important perspective on the quality of healthcare provided. The POA flag can serve as an important information when elaborating sex differences in outcome for KD.

The POA indicator is dependent on the prevalence of the underlying conditions and diagnoses and the associated coding guidelines to a certain ICD code (40). Therefore, we relied on a data-driven approach to assign diagnoses of AKI, CKD and acute-on-chronic KD with the help of a complex rule engine implementing the KDIGO criteria for diagnosis and staging (25,26). Since 2017, based on the official coding rules, AKI and CKD have been coded according to the KDIGO guidelines in Switzerland (40), but documentation of the exact KDIGO staging is often missing in the electronic health records (EHR) and coding and documentation quality remains to be an issue (41).

The aim of this retrospective observational study is to analyse sex differences in pre-existent and hospital acquired AKI with regard to administrative and clinical indicators using the POA flag.

## Materials and Methods

### Design

Routinely collected health data of inpatients in acute care was used in this retrospective observational study. The coding was conducted centralized. The coding of AKI was supported by a highly automated exact and validated calculation of AKI stages.

### Setting

In Switzerland, the Insel Group’s two facilities provide all levels of both in- and outpatient care and contains a tertiary care at the University Hospital Bern (one of the five University Hospital in Switzerland) and four sites with primary and secondary care including specialized care in certain disciplines. Patients show a high level of complexity and many are transferred from other hospitals with complications at admission.

### Study population

The study population included all inpatients of acute care at the Insel Group’s sites from 01.01.2019 to 31.12.2020. Patients at admission being 16 years of age or older were included. No further exclusion criteria were applied.

### Data sources

The following datasets were used: i) Cost data containing information on discharge unit and readmission; ii) Patient Clinical Complexity Level (PCCL) data containing severity scores per case and per diagnosis; iii) POA data containing POA values for selected ICD-10 GM codes per case; iv) Movement data containing the case related movements within the hospital during inpatient stay; v) Medical statistic datasets containing demographics, diagnosis, admission and discharge information per case. All the datasets were merged by using the inpatient case number serving as unique identification (caseID). Cases without POA values and cases missing in cost or movement datasets were deleted. Cases without PCCL values were kept, since the individual diagnosis related CCL values were used to calculate the PCCL. The approach used was (39):

i) Order CCL scores from biggest to smallest for each case; ii) Add a column for row number starting to 1; iii) Calculate *CCL*e* ^*−(row−1)∗0*.*4*^ for each row; iv) Sum all the above values per case;

v) Calculate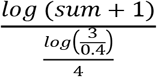; and vi) Round result to get 0, 1, 2, 3, or 4 (values higher than 4 are forced to 4). The data were de-identified from the start and any sensitive information (such as patients’ names or date of birth) were deleted.

## Variables

### Overview

Several variables were used for the current analysis. Details on each variable are presented in the next section. A short overview is provided in Table 1.

**Table 1.**
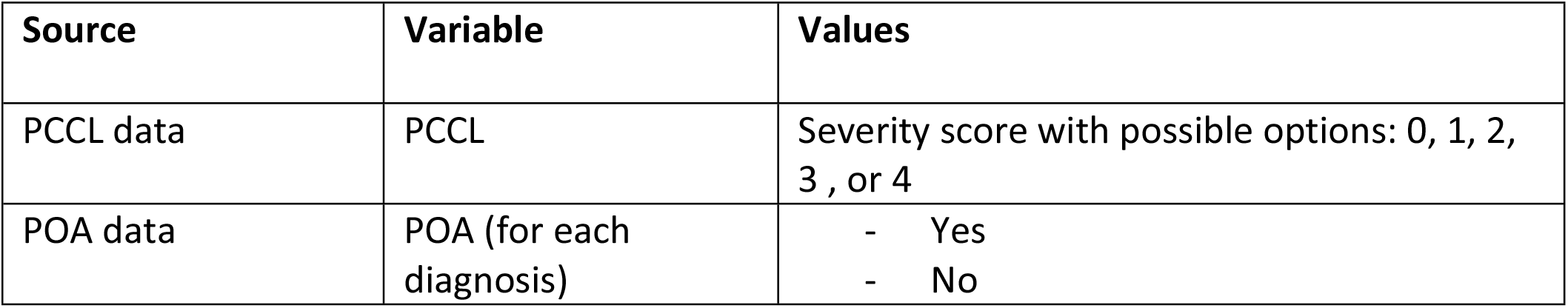
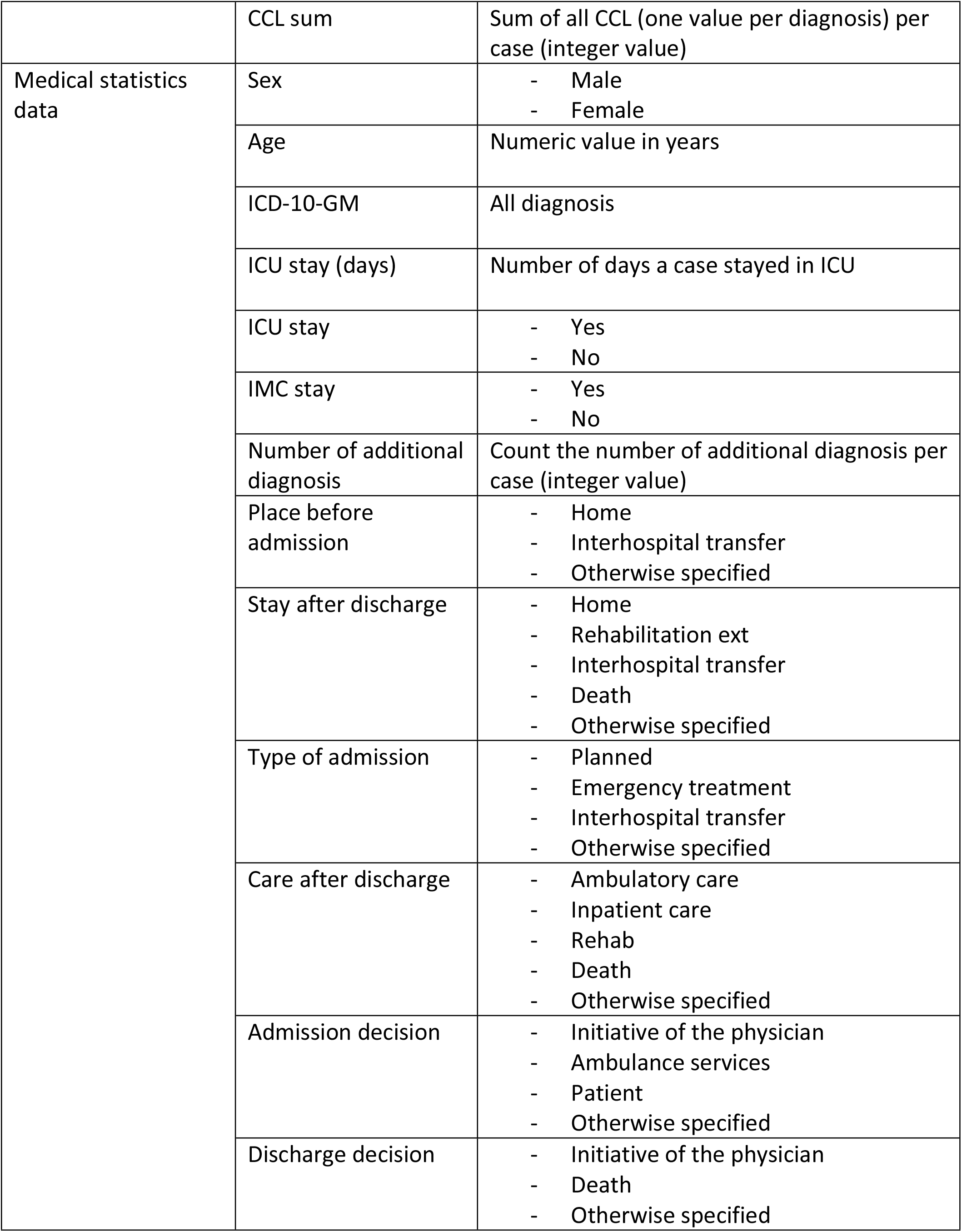
Overview variables.

| Source | Variable | Values |
| --- | --- | --- |
| PCCL data | PCCL | Severity score with possible options: 0, 1, 2, 3, or 4 |
| POA data | POA (for each diagnosis) | - Yes<br>- No |

|  | CCL sum | Sum of all CCL (one value per diagnosis) per case (integer value) |
| --- | --- | --- |
| Medical statistics data | Sex | - Male<br>- Female |
|  | Age | Numeric value in years |
|  | ICD-10-GM | All diagnosis |
|  | ICU stay (days) | Number of days a case stayed in ICU |
|  | ICU stay | - Yes<br>- No |
|  | IMC stay | - Yes<br>- No |
|  | Number of additional diagnosis | Count the number of additional diagnosis per case (integer value) |
|  | Place before admission | - Home<br>- Interhospital transfer<br>- Otherwise specified |
|  | Stay after discharge | - Home<br>- Rehabilitation ext<br>- Interhospital transfer<br>- Death<br>- Otherwise specified |
|  | Type of admission | - Planned<br>- Emergency treatment<br>- Interhospital transfer<br>- Otherwise specified |
|  | Care after discharge | - Ambulatory care<br>- Inpatient care<br>- Rehab<br>- Death<br>- Otherwise specified |
|  | Admission decision | - Initiative of the physician<br>- Ambulance services<br>- Patient<br>- Otherwise specified |
|  | Discharge decision | - Initiative of the physician<br>- Death<br>- Otherwise specified |

### POA indicator

POA reporting was introduced into the standard coding process in 2018. The inclusion list of diagnoses to be reported was specifically designed by the coding department at the Insel Group. It is based on the Hospital Acquired Complications list published by The Australian Commission on Safety and Quality in Health Care (42). The technical implementation into the coding workplace (ID Diacos and SAP-ISH (43,44)) took place in 2017.

The POA coding adhered to the CDA’s ICD-10-CM Official Guidelines for Coding and Reporting, fiscal year 2017, Appendix I (45). Captured values were: 1) Yes = Diagnosis was present at time of inpatient admission; 2) No = Diagnosis was not present at time of inpatient admission; 3) U (unknown) = Documentation insufficient to determine if the condition was present at the time of inpatient admission; 4) W (clinically undetermined) = Provider unable to clinically determine whether the condition was present at the time of inpatient admission; and 5) NA (not assigned) = blank.

For this study, we collapsed the different POA categories into two: 1) “POA no” referring to health care associated conditions and “POA yes” (including yes, W, NA, and U) referring to conditions present at admission(45).

### Sex

In alignment with the WHO and the reported data in the hospitals’ administrative system, we defined the variable sex as the “biological category based on reproductive, anatomical, and genetic characteristics, generally defined as male, female, and intersex” (46). The definition of the term “gender” as given by the WHO does not apply to this study. For the current study, only the answer options “male” and “female” were available, values for biological intersex were not captured.

### ICD codes

The International Statistical Classification Of Diseases And Related Health Problems, 10th revision, German Modification (ICD-10-GM)(47) is the official classification for encoding diagnoses in inpatient care in Switzerland(48).

In this study, the ICD-10 GM coding of category N17 (AKI and stages) relies on a data-driven approach using a complex rule engine (49). Real-time and retrospective data from the hospital’s clinical data warehouse based on a SQL algorithm was used to apply the specific AKI codes to inpatient stays. They were used in this study to define the AKI groups 1-3 and 9 (stages I – III and not classified), see S1 Table for the selection of ICD-10 GM codes used for grouping cases according to the AKI stages.

### Severity indicators CCL and PCCL

In order to outline the possible effects of secondary diagnoses on resource consumption, the ICD-10 GM codes are assessed as a complication or comorbidity for the DRG system. The severity value Complication and Comorbidity Level (CCL) is assigned. A complex algorithm of the SwissDRG system calculates the inpatient case’s specific Patient Clinical Complexity Level (PCCL) from the CCLs (cumulative effect)(39). Neither in the SwissDRG tariff system nor in this study does the calculation of the PCCL and of the CCL level per diagnosis consider whether a condition arose during the episode of care or not.The PCCL values as well as the CCL values summed per case were used in this this study (39).

### Morbidity indicators

Outcome related variables like ICU and IMC stay, inter-hospital transfer and morbidity were selected, see S2 Table. Variables representing morbidity like duration of mechanical ventilation, scores used to quantify, evaluate and allocate nursing workload at intensive care unit level “Nine equivalents of nursing manpower use score” (NEMS) (19) and the severity score “Simplified Acute Physiology Score” (SAPS) (20) were chosen (S2 Table).

### Further variables

Age at admission was also included. Information about admission and discharge was used and when resulting in answer options below 1% were grouped under “otherwise specified”. The variables place before admission, stay after discharge, type of admission, care after discharge, admission decision, discharge decision were included.

### Data analysis

The statistical program R, version 4.2.1 for Linux was used with the following packages: dplyr, reshape2, tidyr, and janitor for data manipulation and tables; stringr for text manipulation ggplot2 and ggpubr for creating and arranging plots, see S3 Table.

In order to describe relevant indicators of morbidity and outcomes a stepwise descriptive analysis of the characteristics was conducted for the study population as a whole and for specific subpopulations (i.e. ICD-10 GM codes). The distribution of the POA values was subsequently stratified for the variable sex. The diagnoses were analysed referring to the hierarchical levels of the ICD-10 GM. The AKI stages required an accumulation of certain 5 digit ICD-10 GM codes (50) per group. The significance of differences in the incidence of health associated diagnosis coding with regard to sex was calculated for all AKI stages. Furthermore, a descriptive analysis of several indicators specified for AKI stage, sex and POA coding was carried out. For reason of small numbers, the coding of acute-on-chronic KD was analysed separately.

Mean with standard deviation and median with interquartile range were computed for numerical variables and percentages for categorical variables. An overview of the population characteristics was conducted. Stratifications for sex and for AKI stages were also conducted. Bar charts were created for stratifications of POA, sex, CKD and/or AKI stages with or without additional variables. Finally, boxplots stratified by AKI staging for numerical variables were performed.

### Ethics

The Ethics Committee of the Canton Bern approved our study KEK-Nr. Req-2018-00328 for the further use of coded, health-related personal data.

## Results

### Final data set

The final datasets included 105’673 inpatient cases and 947’500 diagnoses. Overall, 3865 cases were excluded because of missing PCCL data. An overview of the merging process is shown in Fig 1.

**Fig 1.**
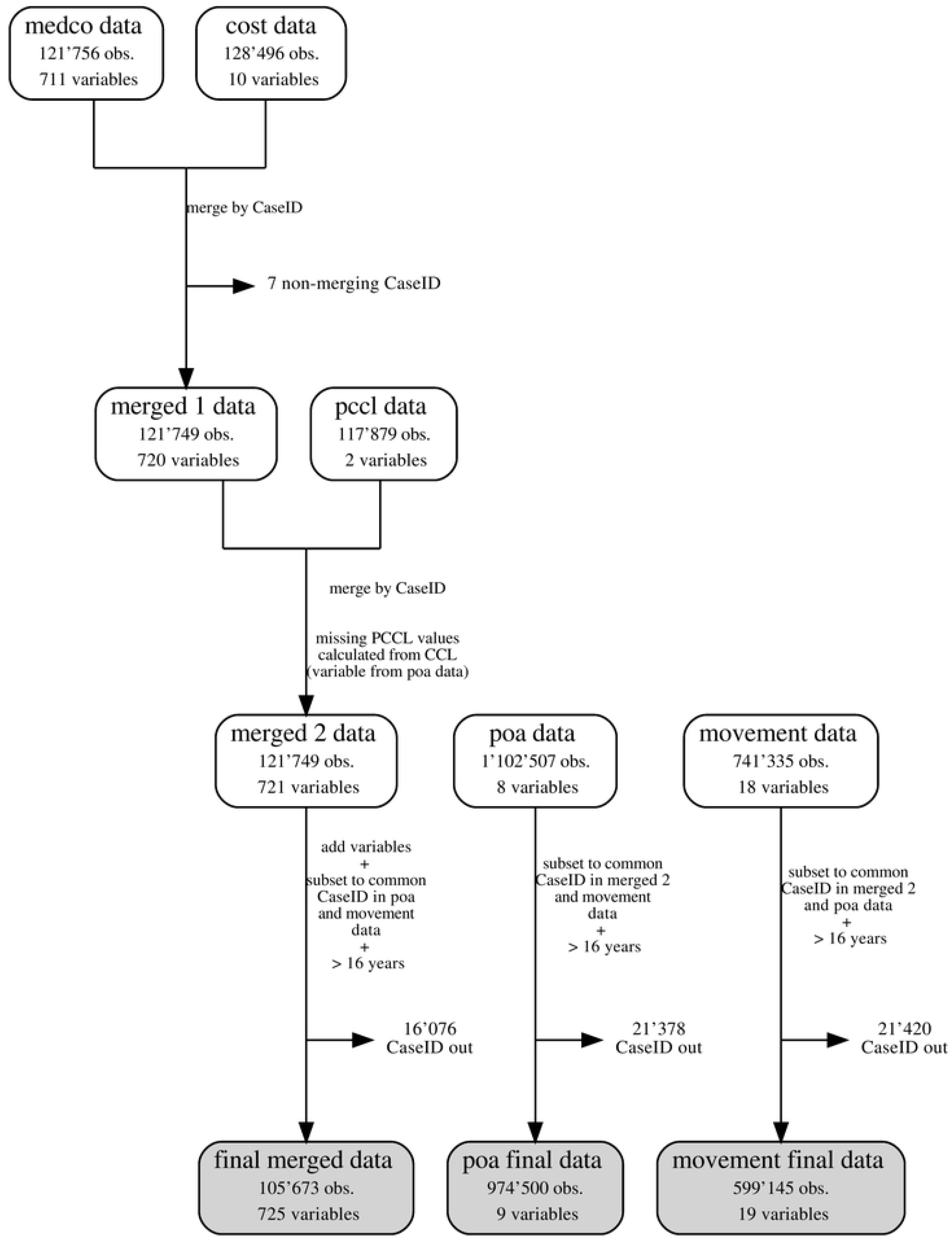
Flowchart data merging.

An overview of the distribution of overview of the population characteristics is given in S4 Fig.

## Stratification by sex

### Overview

Comparing the proportion of POA values associated to indicators of patient history several sex differences can be demonstrated (S5 Fig), e.g. health care associated diagnoses of the male subpopulation show a higher proportion of related cases with interhospital transfer or ICU stay than of the female subpopulation (S6 Table).

### ICD hierarchy

Comparing the ICD blocks, categories and subcategories of ICD-10 GM codes defined for KD the proportion of health care associated diagnoses of all diagnoses of that group is higher in male than in female patients (S7 Table).

Table 2 highlights significant sex differences of health care associated diagnoses for all ICD diagnoses, but not for AKI.

**Table 2.** ICD present on admission (POA yes) and health care associated (POA no) all diagnoses and AKI by sex.

|  | POA no | All diagnoses<br>POA yes | All | POA no | AKI<br>POA yes | All |
| --- | --- | --- | --- | --- | --- | --- |
| Female | 25,747 (5.5%) | 442,364 (94.5%) | 468,111 | 448 (13.6%) | 2,846 (86.4%) | 3,294 |
| Male | 34,090 (5.9%) | 535,711 (94%) | 569,801 | 729 (14.6%) | 4,269 (85.4%) | 4,998 |
| | $\chi^2 = 810860$, df = 1, p-value < 2.2e-16 | | | $\chi^2 = 1.50$, df = 1, p-value = 0.2202 | | |

Analysing exclusively the clinical indicators for the ICD-10 GM chapter N and for block N1 cases related to health care associated diagnoses of male patients show higher values compared to female patients (n diagnoses, SAPS, NEMS, CCL_sum) (S8 and S9 Table).

### ICD 17 AKI stages

8289 ICD diagnoses with POA reporting are grouped into 4024 diagnoses for AKI I, 1340 AKI II and 1412 AKI III. 1345 AKI ICD diagnoses are not classified by AKI stage. The absolute numbers of ICD codes associated to males is higher in all subgroups. The same accounts for the group of acute-on-chronic KD (ICD codes N17 and N18 combined). Nevertheless, only slight differences can be observed in both female and male patients of all AKI and acute-on-chronic diagnoses of the specific subgroups (Fig 2 and S10 and S11 Table).

**Fig 2.**
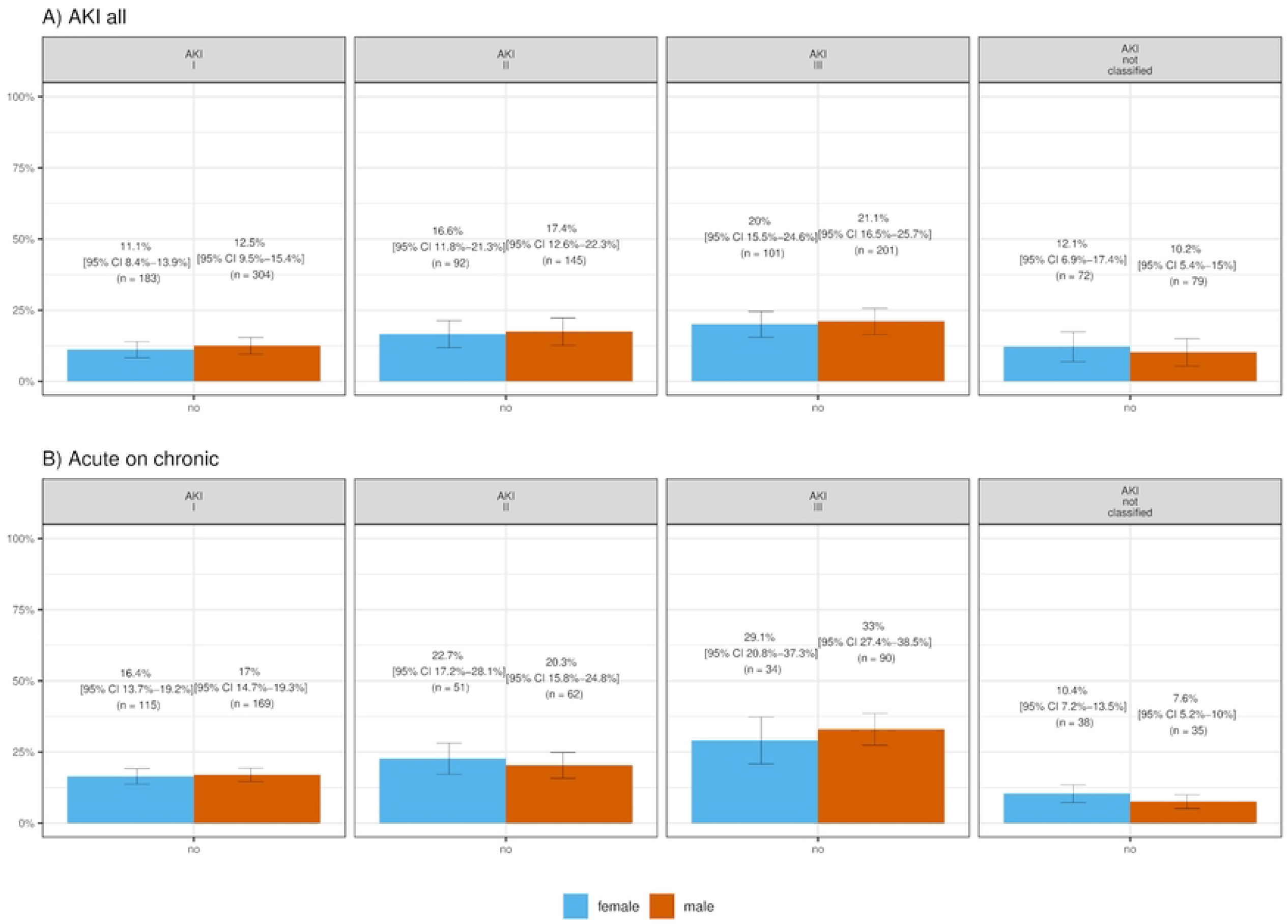
ICD 17 Health care associated AKI and acute-on-chronic staging distribution (POA no) by sex.

Moreover, no significant difference in distribution of health associated diagnoses between female and male cases can be observed in any specific AKI stage. The distribution of AKI and CKD stages for ICD coding of health care associated acute-on-chronic KD (ICD-10 GM N17 and N18) is displayed in Fig 3 and S12 Table.

**Fig 3.**
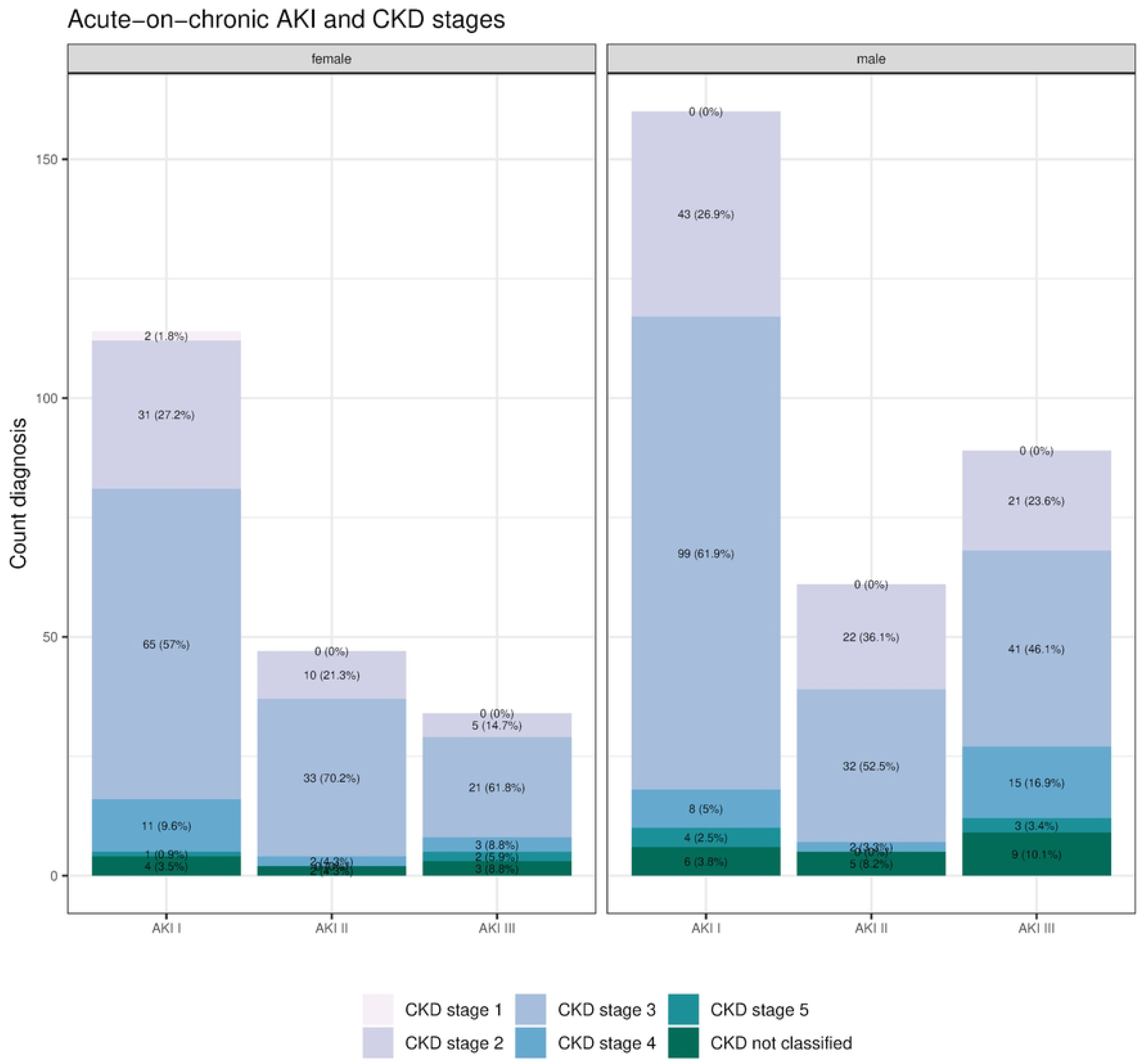
AKI and CKD staging of health care associated acute on chronic KD (POA no) by sex.

With 238 health care associated ICD diagnoses of females and 356 of males respectively the biggest differences between both sex in absolute numbers can be shown for groups AKI I CKD 2 and AKI III CKD 2 and 3. As most of the groups are of small number, we did not execute further analyses.

Figure 4 highlights the proportion of health care associated diagnoses related to the patient history for female and male patients for AKI stages I-III (Fig 4 and S13 Table).

**Fig 4.**
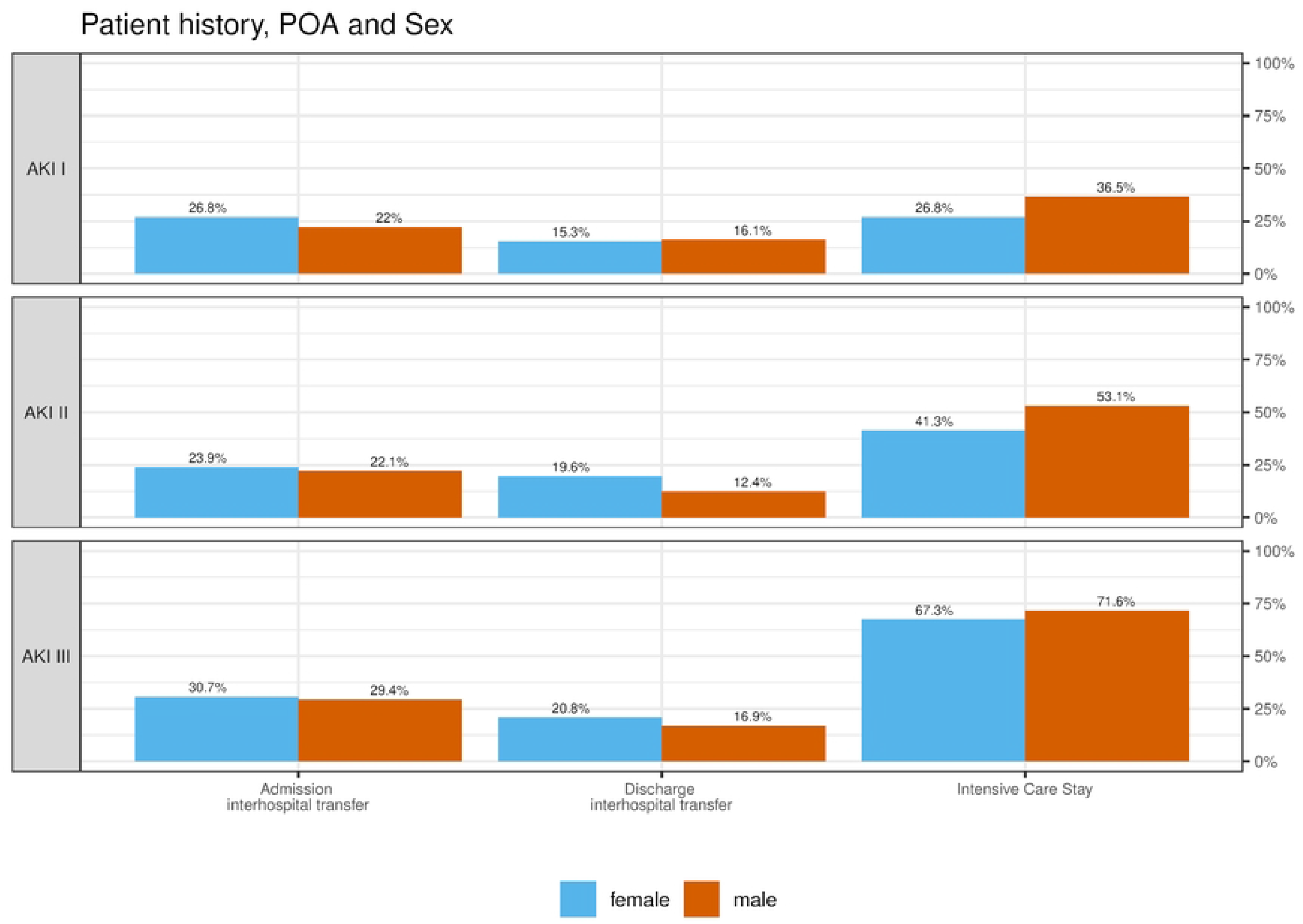
ICD N17 stages of health care associated AKI (POA no) patient history by sex.

The proportion of health care associated diagnoses of females is higher for all AKI stages with regard to “admission – interhospital transfer” and for AKI II and III with regard to “discharge – interhospital transfer” than of males. However, for all AKI stages the proportion of “intensive care” stays is lower.

Compared to males the following differences of the morbidity indicators of females for health care associated AKI can be highlighted:

Significant differences of median values can be demonstrated in AKI I for a shorter duration of ICU stay, in AKI II for a higher age, lower SAPS score and less CCL sum, in AKI III for a higher age and fewer ICD coded diagnoses (Fig 5 and S13).

**Fig 5.**
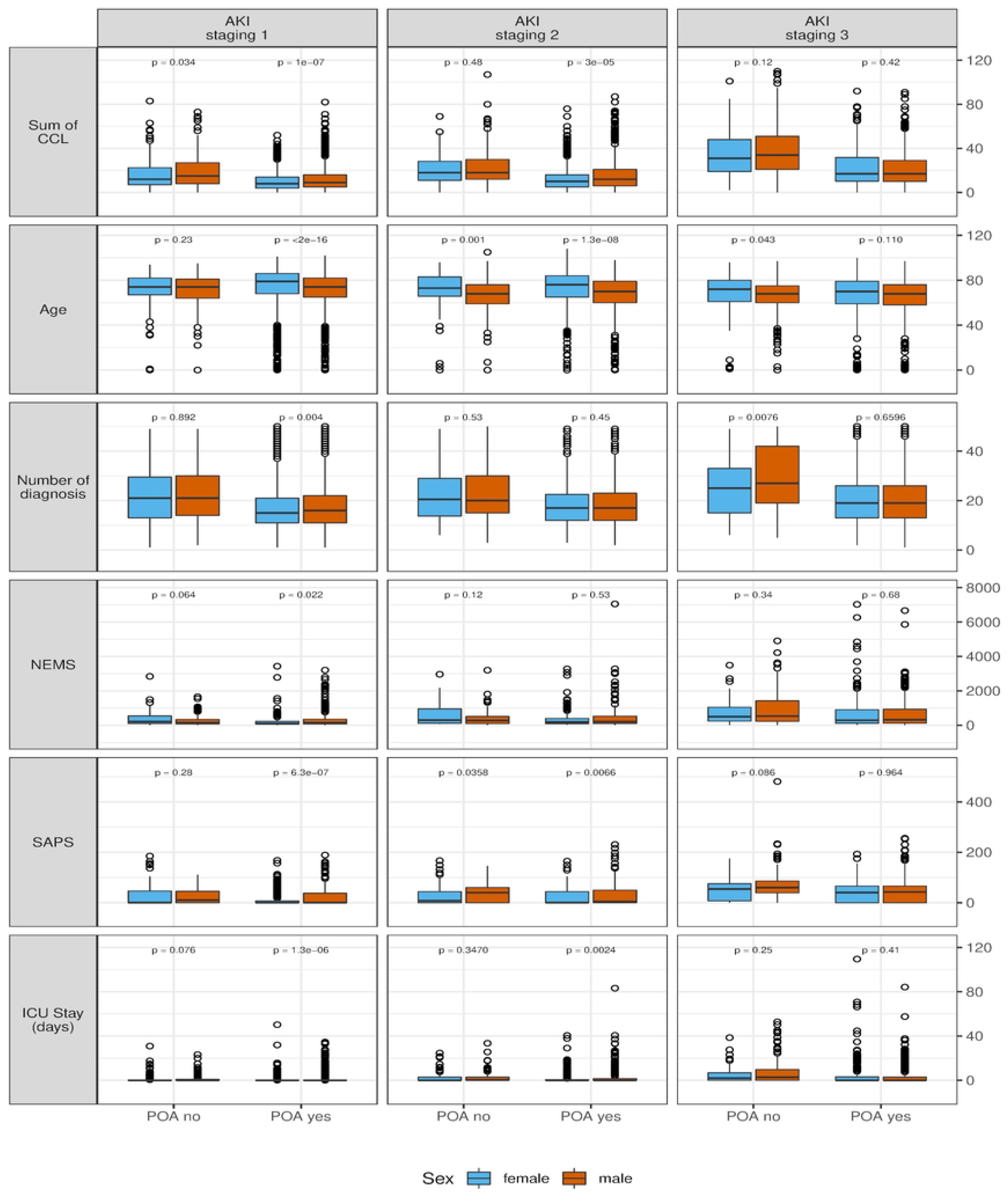
ICD N17 present on admission (POA yes) and health care associated (POA no) AKI morbidity indicators by sex.

### Mortality

The mortality related to health care associated diagnoses of AKI is higher for female patients (all AKI diagnoses 19.3 %, AKI stage II 26.1 %, III 41.6% and acute-on-chronic KD 14.3%, male patients 16.4%, 15.9%, 33.8% and 11.8% respectively) (Fig 6 and S14 Table).

**Fig 6.**
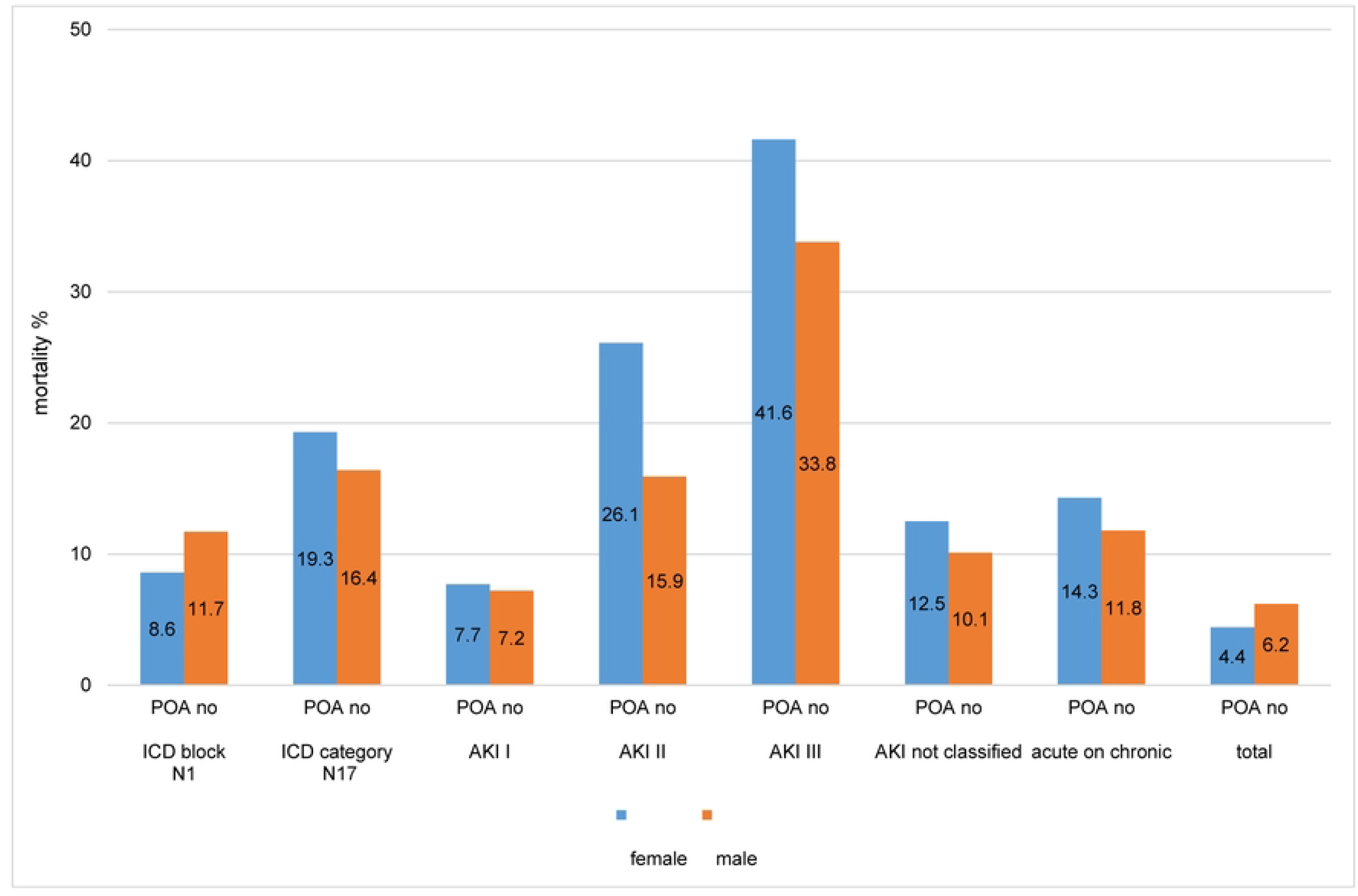
Mortality ICD hierarchy health care associated AKI (POA no) by sex.

## Discussion

### General remarks

The objective of the study was to explore sex differences in the distribution of POA status, of associated patient history and clinical indicators. When analysing subgroups of the final dataset we focused on ICD chapter N “Diseases of the genitourinary system” containing clinically highly relevant diagnoses for both sex and showing significant differences in POA reporting. In this study we could make differences between female and male patients apparent which might induce a deeper analysis of larger populations in future. We could relate the defined outcome indicators to health care associated AKI by making use of the POA reporting and highlight sex differences. The diagnosis-onset reporting using the present on admission indicator was essential to uncover sex differences in health care associated acute kidney injury.

### Main findings

The reporting of health care associated diagnoses corresponds to the defined indicators of patient history and morbidity.

For ICD chapter N “Diseases of the genitourinary system” we found a significant sex difference in the distribution of health care associated diagnoses (females 5.9%, males 5.2%).

The acuity of AKI when health care associated corresponds to patient history e.g. a higher proportion of ICU stays and interhospital transfers. The female population shows a lower proportion of received ICU treatment but higher rates of interhospital transfers. The selected variables of patient history and morbidity were inconsistently linked to the severity of the AKI stages and did not correspond to mortality.

Significant sex differences can be demonstrated for a higher age in health care associated AKI II and III, a shorter duration of ICU stay in AKI I, lower SAPS score in AKI II and III, less CCL sum in AKI I and fewer ICD coded diagnoses in AKI III.

The mortality during hospitalization related to the health care associated diagnoses of all analysed ICD blocks, categories, AKI stages and acute-on-chronic KD stages was higher than of diagnoses of these groups reported not to be health care associated. The mortality was highest for the subgroups of diagnoses AKI II and III and acute-on-chronic KD of female patients (26.1%, 41.6% and 14.3% of health care associated diagnoses related to cases of female patients).

### Discussion of the findings

In this study, we demonstrated the usability of diagnosis-onset reporting and identified the association of post-admission diagnoses with patient history, morbidity, mortality, and sex. However, not all the analyzed groups showed statistically significant differences.

The diagnoses reported to be health care associated relate to a higher proportion of interhospital transfer at admission and discharge, ICU and IMC stays, elevated SAPS and NEMS scores and mechanical ventilation hours, higher age, more diagnoses, a higher CCL sum and a higher PCCL. These findings reflect the clinical situation of complications and comorbidities evolving after admission in inpatient care. Except for older age, the results of the reported health care-associated diagnoses displayed a higher morbidity and more complex patient history related to a higher degree to males than to females. We demonstrated this for all examined diagnoses (AKI and stages, acute-on-chronic KD) and found statistically significant differences in some subgroups between the POA values and sex. In our population the proportion of the diagnoses related to female patients was lower than related to male patients. In general, a low detection rate of diagnoses and stages may cause a lower admission rate to the ICU (15,17,51) and underestimated morbidity and conservative treatment could lead to worse outcome (52,53). A lower detection rate can be discussed but does not apply for AKI, CKD and acute-on-chronic KD at our institution as by automated detection (CDSS) all diagnoses and stages are captured during inpatient stay. However, the CDSS is currently not implemented for clinical use into the information system. Therefore, the calculated diagnoses and stages do not have an immediate effect on the patients’ treatment if not recognized otherwise during inpatient stay. Nevertheless, the diagnoses we analysed in this study were approved by the clinician before effectively coded. We could prove in a former study that this method supports the process and reliability of diagnosis (49).

Our results of the analysis of the AKI stages showed lower values for indicators related to morbidity in the group of diagnoses associated to females, but a higher mortality. Therefore, a deeper evaluation of the implied sex differences in staging of KD should follow.

In past studies it could be shown that the incidence of HA-AKI was greater in males than females when KDIGO criteria were used but differences disappeared when staging was conducted according to the RIFLE criteria (27). On the other hand we assume that care seeking behaviour and the offered intensity of care might be influenced by the patient’s age alone indipendently from comorbidities (7,53,54). Both could explain the higher mortality we observed in the female patients.

As AKI is not an independent diagnosis but expression of a heterogeneous set of disorders characterized by an acute organ dysfunktion, the diagnosis-onset reporting and sex differences need further investigation with regard to risk adjustment. Observational studies have shown a variety of risk factors like pre-existing susceptibilities of individual patients related to the severity of AKI (55). Concerning risk adjustement and definition of outcome indicators used in this and in other studies it should be mentioned that the underlying data might not appropriately represent females. For instance, neither the validation of the NEMS nor of the SAPS scores (and as preliminary work of the Therapeutic Intervention Scoring System -TISS-28) was based on equally distributed datasets of female and male ICU patients (19–21). Therefore, it can be questioned, if they are as appropriate to assess morbidity / serverity and nursing workload for female patients as they are for male. Furthermore, risk adjustment for hypertension, diabetes mellitus, cardiological complications and liver cirrhosis which often accompany AKI, should consider that these conditions are not distributed equally and related research is impacted by lacking sex stratification (10,56).

As mortality in our study was highest for the subgroups of health care associated diagnoses AKI II and III and acute-on-chronic KD of female patients we should expect matching results with regard to indicators for patient history and morbidity, which we did only partially. As the only unambiguous measure was mortality, this may question the appropriateness of the other chosen indicators. The mortality rate related to the diagnoses may even be higher than demonstrated here because we only considered in-hospital mortality. To impove the future work at our institution the integration of e.g. the Charlson Comobidity Index (57) into the health record would facilitate risk adjustment. However, it could be demonstrated in a thesis (ICD-10 GM codes; German dataset) that the the rather simple approach of counting the number of coded ICD codes (58), which we applied in our study, is a usable severity indicator. Due to the small numbers, we reported only a few results for acute-on-chronic KD. Essentially they agree in interpretation with the results of AKI and staging, as we observed a higher mortality. Also, patient history and morbidity show higher values and proportions for the health care associated conditions and for mortality in the group of diagnoses associated to female patients. Nevertheless, the staging of AKI and CKD in acute-on-chronic KD should be analysed further, as until now it is often underreported at discharge and administrative data is not available to the extent of AKI and CKD at our institution and elsewhere (36)(59). Therefore, not only the diagnosis-timing of the highest stage should be analysed, but the disease progression during hospitalization measured by encoding all stages. For this purpose, instead of a simple POA flag a time stamp for the ICD codes would be suitable. As the CDSS is also designed to monitor AKI progression, it encourages analyses of the course of KD in future.

Our analysis shows that by implementing the POA indicator in health care systems worldwide the utility of routinely collected health data could be improved, especially for risk adjustment. National stakeholders show a high interest in quality monitoring and benchmarking by further developed administrative datasets (60) and in improved risk models. A proposal for extension of the national data set for diagnosis timing was put forward by the authors of this study to the Swiss Federal Statistic Office in 2018 and was rejected (61). Only on regional level the Canton Zurich will introduce POA reporting in 2023 (62).

Based on the findings we suggest that the POA indicators should be considered for medical statistics and quality management (63,64).

### Clinical significance

The results show that POA reporting is an important contribution to make patient groups with high risk for worse outcomes transparent (mortality as a hard indicator). To improve risk adjusted analyses, monitoring, and treatment the POA reporting delivers important information. Moreover, in the specific context of AKI the results show the need to monitor progression of disease closely and to stratify patient groups for differentiation.

### Strength

To our knowledge, this is the first study to analyse the association of the POA indicator and sex differences to outcome and morbidity data using a highly reliable and precise dataset of ICD coding of AKI with an exact staging and of acute-on-chronic KD (according to KDIGO). A data-driven automatic approach including pre-hopitalization laboratory values ensures the reliable diagnoses, staging and subsequent ICD coding (49). Therefore, in this study data is not impacted by a low ICD coding and / or documentation quality of the diagnoses themselves.

Available data of ICD coding of acute-on-chronic disease based on routinely collected health data is a strength as until now the diagnosis acute-on-chronic KD is poorly understood and very often underreported in documentation, ICD coding and POA reporting (36,41).

### Limitations

As we started POA reporting at our hospitals only in 2018, data of 2 fiscal years is availabe until now, which limits the population size of the current analyses. Due to small numbers it was not possible to interpret sufficiently significance levels of some of the subgroups. Moreover, with regard to mortality we were limited to in-hospital mortality and did not consider mortality after hospitalization, although it is known to be elevated in association with KD (65). Another drawback of this retrospective observational study is the lacking risk adjustment, especially for relevant comorbidities and age, which might contribute to sex differences. This study does not cover overall incidence and prevalence of KD. Due to Swiss national and cantonal legislation, sex is captured with the values either “female” or “male” in administrative systems (66) and the medical statistic dataset of the FSO (variable 1.1.V01: 1-Mann, 2-Frau)(38). Therefore, the data lacks further differentiation in this study.

## Conclusions

The POA indicator proofed to be a valuable variable to flag diagnoses arising during the episode of care for analysis of administrative data. Diagnoses of AKI and acute-on-chronic KD could be successfully indexed and associated to patient history, morbidity, mortality and sex. Sex differences of health care associated AKI and stages could be made transparent. The higher mortality of female patients with health care associated diagnoses could not be associated to a higher morbidity of that group as defined in this study. Ruling out the possibility of sex related underdiagnosis of AKI staging, we recommed the implementation of the automated algorithm to assign the diagnoses of CKD and AKI as a CDSS into production for clinical use. Moreover, a deeper evaluation of the implied sex differences in staging of KD according to different criteria is needed.

In order to obtain a larger study population especially for the diagnosis acute-on-chronic disease the neccessity of a diagnosis-onset reporting in national health statistic can be confirmed. Awareness of biases and disparities by transparency is a sensible step in improving clinical decision-making.

## Data Availability

All relevant data are within the manuscript and its Supporting Information files.

## Acknowledgments

We would like to thank Agnes Schöpfer for ICD code mapping and our colleagues from the Insel Gruppe Medizincontrolling Coding Team for their tireless efforts in tracking down all the individual codes. ID Diacos and SAP-ISH for technical support.

## Supporting information captions

S1 Table. Groups AKI ICD 17 5 digit.

S2 Table. Variables morbidity indicators and patient history. S3 Table. Software packages.

S4 Fig. Overview of the population characteristics.

S5 Fig. Patient history female male overview.

S6 Table. Patient history female male overview.

S7 Table. ICD chapter N health care associated (POA no) distribution by sex.

S8 Table. ICD all 2 digit level health care associated (POA no) morbidity indicators by sex.

S9 Table. ICD N 1 and 2 digit level health care associated (POA no) morbidity indicators by sex.

S10 Table. ICD 17 AKI staging significance health care associated (POA no) no by sex.

S11 Table. AKI and CKD staging of health care associated acute on chronic KD (POA no) by sex.

S12 Table. ICD N17 AKI 5 digit level health care associated (POA no) patient history by sex.

S13 Table. ICD N17 AKI health care associated (POA no) morbidity indicators by sex.

S14 Table. Mortality ICD hierarchy health care associated (POA no) by sex.

## References

1. Merkatz RB. Inclusion of Women in Clinical Trials: A Historical Overview of Scientific Ethical and Legal Issues. J Obstet Gynecol Neonatal Nurs [Internet]. 1998;27(1):78–84. Available from: https://www.sciencedirect.com/science/article/pii/S0884217515335267

2. Fullerton JT, Sadler GR. Ethical considerations related to the inclusion of women in clinical trials. J Midwifery Womens Health [Internet]. 2004;49(3):194–202. Available from: https://www.sciencedirect.com/science/article/pii/S152695230300429X

3. Polit DF, Beck CT. Generalization in quantitative and qualitative research: Myths and strategies. Int J Nurs Stud [Internet]. 2010;47(11):1451–8. Available from: https://www.sciencedirect.com/science/article/pii/S0020748910002063

4. Franconi F, Campesi I, Colombo D, Antonini P. Sex-Gender Variable: Methodological Recommendations for Increasing Scientific Value of Clinical Studies. Cells. 2019 May;8(5).

5. U.S. Department of Health and Human Services National Institutes of Health. NIH Policy and Guidelines on The Inclusion of Women and Minorities as Subjects in Clinical Research [Internet]. 2017. Available from: https://grants.nih.gov/policy/inclusion/women-and-minorities.htm

6. Roth GA, Abate D, Abate KH, Abay SM, Abbafati C, Abbasi N, et al. Global, regional, and national age-sex-specific mortality for 282 causes of death in 195 countries and territories, 1980–2017: a systematic analysis for the Global Burden of Disease Study 2017. Lancet [Internet]. 2018;392(10159):1736–88. Available from: https://www.sciencedirect.com/science/article/pii/S0140673618322037

7. Spector ND, Overholser B. Examining Gender Disparity in Medicine and Setting a Course Forward. JAMA Netw Open [Internet]. 2019 Jun 28;2(6):e196484–e196484. Available from: https://doi.org/10.1001/jamanetworkopen.2019.6484

8. Verdonk P, Benschop YWM, de Haes HCJM, Lagro-Janssen TLM. From gender bias to gender awareness in medical education. Adv Heal Sci Educ [Internet]. 2009;14(1):135–52. Available from: https://doi.org/10.1007/s10459-008-9100-z

9. Thompson AE, Anisimowicz Y, Miedema B, Hogg W, Wodchis WP, Aubrey-Bassler K. The influence of gender and other patient characteristics on health care-seeking behaviour: a QUALICOPC study. BMC Fam Pract [Internet]. 2016;17(1):38. Available from: https://doi.org/10.1186/s12875-016-0440-0

10. Feldman S, Ammar W, Lo K, Trepman E, van Zuylen M, Etzioni O. Quantifying Sex Bias in Clinical Studies at Scale With Automated Data Extraction. JAMA Netw Open [Internet]. 2019 Jul 3;2(7):e196700–e196700. Available from: https://doi.org/10.1001/jamanetworkopen.2019.6700

11. Thurlow JS, Joshi M, Yan G, Norris KC, Agodoa LY, Yuan CM, et al. Global Epidemiology of End-Stage Kidney Disease and Disparities in Kidney Replacement Therapy. Am J Nephrol. 2021;52(2):98–107.

12. Elgohary M, Saudy A. Global Prevalence of Chronic Kidney Disease. 21st Int Conf Struct Mech React Technol (SMiRT 21). 2011;1–18.

13. Bikbov B, Perico N, Remuzzi G. Disparities in Chronic Kidney Disease Prevalence among Males and Females in 195 Countries: Analysis of the Global Burden of Disease 2016 Study. Nephron [Internet]. 2018;313–8. Available from: https://www.karger.com/DOI/10.1159/000489897

14. Carrero JJ, Hecking M, Chesnaye NC, Jager KJ. Sex and gender disparities in the epidemiology and outcomes of chronic kidney disease. Nat Rev Nephrol [Internet]. 2018;14(3):151–64. Available from: https://doi.org/10.1038/nrneph.2017.181

15. Valentin A, Jordan B, Lang T, Hiesmayr M, Metnitz PGH. Gender-related differences in intensive care: A multiple-center cohort study of therapeutic interventions and outcome in critically ill patients*. Crit Care Med [Internet]. 2003;31(7). Available from: https://journals.lww.com/ccmjournal/Fulltext/2003/07000/Gender_related_differences_in_intensive_careA.2.aspx

16. Carrero J-J, Hecking M, Ulasi I, Sola L, Thomas B. Chronic Kidney Disease, Gender, and Access to Care: A Global Perspective. Semin Nephrol [Internet]. 2017;37(3):296–308. Available from: https://www.sciencedirect.com/science/article/pii/S0270929517300098

17. Hill A, Ramsey C, Dodek P, Kozek J, Fransoo R, Fowler R, et al. Examining mechanisms for gender differences in admission to intensive care units. Health Serv Res [Internet]. 2020;55(1):35–43. Available from: https://onlinelibrary.wiley.com/doi/abs/10.1111/1475-6773.13215

18. Portoles JM, Lopez P, De Valdenebro Recio M, Sanchez Briales P, Luisa Serrano Salazar M, Ramos-Vegue A, et al. MO331: Acute Kidney Failure as a Risk Factor for Longer Hospital Stay, Renal Replacement Therapy and Mortality: A Public Health System Analysis. Nephrol Dial Transplant [Internet]. 2022 May 1;37(Supplement_3):gfac068.041. Available from: https://doi.org/10.1093/ndt/gfac068.041

19. Miranda DR, Moreno R, Iapichino G. Nine equivalents of nursing manpower use score (NEMS). Intensive Care Med [Internet]. 1997;23(7):760–5. Available from: https://doi.org/10.1007/s001340050406

20. Le Gall J-R, Lemeshow S, Saulnier F. A New Simplified Acute Physiology Score (SAPS II) Based on a European/North American Multicenter Study. JAMA [Internet]. 1993 Dec 22;270(24):2957–63. Available from: https://doi.org/10.1001/jama.1993.03510240069035

21. Reis Miranda D, de Rijk A, Schaufeli W. Simplified Therapeutic Intervention Scoring System: The TISS-28 items--Results from a multicenter study. Crit Care Med [Internet]. 1996;24(1). Available from: https://journals.lww.com/ccmjournal/Fulltext/1996/01000/Simplified_Therapeutic_Intervention_Scoring.12.aspx

22. Cobo G, Hecking M, Port FK, Exner I, Lindholm B, Stenvinkel P, et al. Sex and gender differences in chronic kidney disease: progression to end-stage renal disease and haemodialysis. Clin Sci [Internet]. 2016 Jun 1;130(14):1147–63. Available from: https://doi.org/10.1042/CS20160047

23. O’Leary JG, Wong F, Reddy KR, Garcia-Tsao G, Kamath PS, Biggins SW, et al. Gender-Specific Differences in Baseline, Peak, and Delta Serum Creatinine: The NACSELD Experience. Dig Dis Sci [Internet]. 2017;62(3):768–76. Available from: https://doi.org/10.1007/s10620-016-4416-7

24. Neugarten J, Golestaneh L. Sex Differences in Acute Kidney Injury. Semin Nephrol [Internet]. 2022;42(2):208–18. Available from: https://www.sciencedirect.com/science/article/pii/S0270929522000183

25. Eknoyan G LN. KDIGO 2012 Clinical Practice Guideline for the Evaluation and Management of Chronic Kidney Disease [Internet]. KDIGO 2012 clinical practice guideline for the evaluation and management of chronic kidney disease. 2012. Available from: https://kdigo.org/guidelines/ckd-evaluation-and-management/

26. Kellum JA, Lameire N, Aspelin P, Barsoum RS, Burdmann EA GS et al. KDIGO Clinical Practice Guideline for Acute Kidney Injury [Internet]. Improving global outcomes (KDIGO) acute kidney injury work group. KDIGO clinical practice guideline for acute kidney injury. 2012. Available from: https://kdigo.org/guidelines/acute-kidney-injury/

27. Srisawat N, Sileanu FE, Murugan R, Bellomo R, Calzavacca P, Cartin-Ceba R, et al. Variation in Risk and Mortality of Acute Kidney Injury in Critically Ill Patients: A Multicenter Study. Am J Nephrol [Internet]. 2015;41(1):81–8. Available from: https://www.karger.com/DOI/10.1159/000371748

28. Lewington AJP, Cerdá J, Mehta RL. Raising awareness of acute kidney injury: a global perspective of a silent killer. Kidney Int [Internet]. 2013;84(3):457–67. Available from: https://www.sciencedirect.com/science/article/pii/S0085253815559917

29. Gaudry S, Hajage D, Benichou N, Chaïbi K, Barbar S, Zarbock A, et al. Delayed versus early initiation of renal replacement therapy for severe acute kidney injury: a systematic review and individual patient data meta-analysis of randomised clinical trials. Lancet [Internet]. 2020 Apr [cited 2020 Apr 24];0(0). Available from: https://linkinghub.elsevier.com/retrieve/pii/S0140673620305316

30. Lima-Posada I, Portas-Cortés C, Pérez-Villalva R, Fontana F, Rodríguez-Romo R, Prieto R, et al. Gender Differences in the Acute Kidney Injury to Chronic Kidney Disease Transition. Sci Rep [Internet]. 2017;7(1):12270. Available from: https://doi.org/10.1038/s41598-017-09630-2

31. Couser WG, Remuzzi G, Mendis S, Tonelli M. The contribution of chronic kidney disease to the global burden of major noncommunicable diseases. Kidney Int [Internet]. 2011 Dec 2 [cited 2019 Aug 9];80(12):1258–70. Available from: https://www.sciencedirect.com/science/article/pii/S0085253815550047?via%3Dihub

32. Jurawan N, Pankhurst T, Ferro C, Nightingale P, Coleman J, Rosser D, et al. Hospital acquired Acute Kidney Injury is associated with increased mortality but not increased readmission rates in a UK acute hospital. BMC Nephrol [Internet]. 2017;18(1):317. Available from: https://doi.org/10.1186/s12882-017-0729-9

33. Wang HE, Muntner P, Chertow GM, Warnock DG. Acute Kidney Injury and Mortality in Hospitalized Patients. Am J Nephrol [Internet]. 2012;35(4):349–55. Available from: https://www.karger.com/DOI/10.1159/000337487

34. Huang L, Xue C, Kuai J, Ruan M, Yang B, Chen X, et al. Clinical Characteristics and Outcomes of Community-Acquired versus Hospital-Acquired Acute Kidney Injury: A Meta-Analysis. Kidney Blood Press Res [Internet]. 2019;44(5):879–96. Available from: https://www.karger.com/DOI/10.1159/000502546

35. Wonnacott A, Meran S, Amphlett B, Talabani B, Phillips A. Epidemiology and Outcomes in Community-Acquired Versus Hospital-Acquired AKI. Clin J Am Soc Nephrol [Internet]. 2014 Jun 6;9(6):1007 LP – 1014. Available from: http://cjasn.asnjournals.org/content/9/6/1007.abstract

36. Hamroun A, Frimat L, Laville M, Metzger M, Combe C, Fouque D, et al. New insights into acute-on-chronic kidney disease in nephrology patients: the CKD-REIN study. Nephrol Dial Transplant [Internet]. 2022 Sep 1;37(9):1700–9. Available from: https://doi.org/10.1093/ndt/gfab249

37. Shlipak MG, Tummalapalli SL, Boulware LE, Grams ME, Ix JH, Jha V, et al. The case for early identification and intervention of chronic kidney disease: conclusions from a Kidney Disease: Improving Global Outcomes (KDIGO) Controversies Conference. Kidney Int [Internet]. 2021 Jan 1 [cited 2022 Apr 13];99(1):34–47. Available from: https://pubmed.ncbi.nlm.nih.gov/33127436/

38. Bundesamt für Statistik (BFS). Medizinische Statistik der Krankenhäuser [Internet]. Medizinische Statistik der Krankenhäuser - Variablen der Medizinischen Statistik. Spezifikationen gültig ab 1.1.2019. 2019 [cited 2019 Jun 12]. Available from: https://www.bfs.admin.ch/bfs/de/home/statistiken/gesundheit/erhebungen/ms.html

39. SwissDRG. SwissDRG System 9.0/2020 [Internet]. 2019 [cited 2022 Jul 15]. Available from: https://www.swissdrg.org/de/akutsomatik/archiv-swissdrg-system/swissdrg-system-902020

40. Bundesamt für Statistik. Kodierungshandbuch. Instrumente zur medizinischen Kodierung. 2020.

41. Vlasschaert MEO, Bejaimal SAD, Hackam DG, Quinn R, Cuerden MS, Oliver MJ, et al. Validity of Administrative Database Coding for Kidney Disease: A Systematic Review. Am J Kidney Dis [Internet]. 2011;57(1):29–43. Available from: https://www.sciencedirect.com/science/article/pii/S0272638610013983

42. Australian Commission on Safety and Quality in Health Care. Hospital-Acquired Complications (HACs) List - Specifications - Version 3.1 (12th edn) [Internet]. 2019 [cited 2022 Jul 15]. Available from: https://www.safetyandquality.gov.au/publications-and-resources/resource-library/hospital-acquired-complications-hacs-list-specifications-version-31-12th-edn

43. SAP. SAP (Schweiz) AG „Systemanalyse Programmentwicklung” [Internet]. Imressum. 2022. Available from: https://www.sap.com/swiss/about/legal/impressum.html

44. ID. ID Suisse AG “Information und Dokumentation im Gesundheitswesen” [Internet]. Impressum. 2022. Available from: https://www.id-suisse-ag.ch/impressum/

45. Health C for M and MS (CMS) and the NC for, (NCHS) S. ICD-10-CM Official Guidelines for Coding and Reporting [Internet]. 2017. 109–114 p. Available from: https://www.cms.gov/medicare/coding/icd10/downloads/2017-icd-10-cm-guidelines.pdf

46. World Health Organization. Sexual health [Internet]. Sexual Health definitions. 2022. Available from: https://www.who.int/health-topics/sexual-health#tab=tab_2

47. Deutsches Institut für Medizinische Dokumentation und Information. ICD-10-GM [Internet]. ICD-10_GM. 2016 [cited 2019 Sep 23]. Available from: https://www.dimdi.de/dynamic/de/klassifikationen/icd/icd-10-gm/

48. Bundesamt für Statistik. Instrumente zur medizinischen Kodierung [Internet]. Gültige Instrumente zur medizinischen Kodierung je Jahr. [cited 2020 Nov 25]. Available from: https://www.bfs.admin.ch/bfs/de/home/statistiken/gesundheit/nomenklaturen/medkk/instrumente-medizinische-kodierung.html

49. Triep K, Leichtle AB, Meister M, Fiedler GM, Endrich O. Real-world Health Data and Precision for the Diagnosis of Acute Kidney Injury, Acute-on-Chronic Kidney Disease, and Chronic Kidney Disease: Observational Study. JMIR Med Inf 2022;10(1)e31356 https://medinform.jmir.org/2022/1/e31356 [Internet]. 2022 Jan 25 [cited 2022 Apr 14];10(1):e31356. Available from: https://medinform.jmir.org/2022/1/e31356

50. Federal Institute for Drugs and Medical Devices Germany. Tabular List of ICD-10-GM [Internet]. Classifications. 2022. Available from: https://www.bfarm.de/EN/Code-systems/Classifications/ICD/ICD-10-GM/Tabular-list/_node.html

51. Ancochea J, Izquierdo JL, Group SC-19 R, Soriano JB. Evidence of gender bias in the diagnosis and management of COVID-19 patients: A Big Data analysis of Electronic Health Records. medRxiv [Internet]. 2020 Jan 1;2020.07.20.20157735. Available from: http://medrxiv.org/content/early/2020/07/26/2020.07.20.20157735.abstract

52. Kister TS, Remmler J, Schmidt M, Federbusch M, Eckelt F, Isermann B, et al. Acute kidney injury and its progression in hospitalized patients-Results from a retrospective multicentre cohort study with a digital decision support system. PLoS One [Internet]. 2021 Jul 1 [cited 2022 Apr 25];16(7). Available from: https://pubmed.ncbi.nlm.nih.gov/34252151/

53. Hansen K. Gender Bias. In: Raz Mandaand Pouryahya P, editor. Decision Making in Emergency Medicine: Biases, Errors and Solutions [Internet]. Singapore: Springer Singapore; 2021. p. 167–72. Available from: https://doi.org/10.1007/978-981-16-0143-9_27

54. Tong A, Evangelidis N, Kurnikowski A, Lewandowski M, Bretschneider P, Oberbauer R, et al. Nephrologists’ Perspectives on Gender Disparities in CKD and Dialysis. Kidney Int Reports [Internet]. 2022;7(3):424–35. Available from: https://www.sciencedirect.com/science/article/pii/S2468024921015059

55. Schiffl H. Gender differences in the susceptibility of hospital-acquired acute kidney injury: more questions than answers. Int Urol Nephrol [Internet]. 2020;52(10):1911–4. Available from: https://doi.org/10.1007/s11255-020-02526-7

56. Weber AM, Gupta R, Abdalla S, Cislaghi B, Meausoone V, Darmstadt GL. Gender-related data missingness, imbalance and bias in global health surveys. BMJ Glob Heal [Internet]. 2021 Nov 1;6(11):e007405. Available from: http://gh.bmj.com/content/6/11/e007405.abstract

57. Charlson ME, Pompei P, Ales KL, MacKenzie CR. A new method of classifying prognostic comorbidity in longitudinal studies: Development and validation. J Chronic Dis. 1987;40(5):373–83.

58. Hagn S. Vergleich verschiedener Komorbiditäts-Scores in Routinedaten der stationären Versorgung [Internet]. Ludwig-Maximilians-Universität München; 2014. Available from: https://edoc.ub.uni-muenchen.de/17118/1/Hagn_Stefan.pdf

59. Siew ED, Basu RK, Wunsch H, Shaw AD, Goldstein SL, Ronco C, et al. Optimizing Administrative Datasets to Examine Acute Kidney Injury in the Era of Big Data: Workgroup Statement from the 15th ADQI Consensus Conference. Can J Kidney Heal Dis [Internet]. 2016 Jan 1;3:98. Available from: https://doi.org/10.1186/s40697-016-0098-5

60. Triep K, Beck T, Donzé J, Endrich O. Diagnostic value and reliability of the present-on-admission indicator in different diagnosis groups: pilot study at a Swiss tertiary care center. BMC Health Serv Res [Internet]. 2019 Dec 9 [cited 2019 Nov 17];19(1):23. Available from: https://bmchealthservres.biomedcentral.com/articles/10.1186/s12913-018-3858-3

61. Bundesamt für Statistik. Prozess zum Antragsverfahren [Internet]. Medizinische Statistik der Krankenhäuser. 2022 [cited 2022 Nov 6]. Available from: https://www.bfs.admin.ch/bfs/de/home/statistiken/gesundheit/nomenklaturen/medkk/antragsverfahren.assetdetail.22604906.html

62. Gesundheit ZGA für. Abgabetermine Daten 2022, Information zu SDEP und KS. 2022;1–10. Available from: https://www.zh.ch/content/dam/zhweb/bilder-dokumente/themen/gesundheit/gesundheitsversorgung/spitaeler_kliniken/daten_und_statistik_der_listenspitaeler/datenerhebung/datenerhebung-2022/julibrief_2022.pdf

63. GQMG. Empfehlungen der GQMG zur Einführung eines Present-On-Admission-Kennzeichens (POA) für administrative Routinedaten in Krankenhäusern. Z Evid Fortbild Qual Gesundhwes [Internet]. 2012;106(3):231–2. Available from: https://www.sciencedirect.com/science/article/pii/S1865921712000797

64. Dalton JE, Glance LG, Mascha EJ, Ehrlinger J, Chamoun N, Sessler DI. Impact of Present-on-admission Indicators on Risk-adjusted Hospital Mortality Measurement. Anesthesiology [Internet]. 2013 Jun 1;118(6):1298–306. Available from: https://doi.org/10.1097/ALN.0b013e31828e12b3

65. Gameiro J, Marques F, Lopes JA. Long-term consequences of acute kidney injury: a narrative review. Clin Kidney J [Internet]. 2020 Nov 19;14(3):789–804. Available from: https://pubmed.ncbi.nlm.nih.gov/33777362

66. Eidgenossenschaft DBS. Geschlecht und Vornamen im Personenstandsregister unbürokratisch ändern [Internet]. Medienmitteilung. 2019. Available from: https://www.ejpd.admin.ch/ejpd/de/home/aktuell/news/2019/2019-12-063.html

